# Clinical and molecular characterization of COVID-19 hospitalized patients

**DOI:** 10.1101/2020.05.22.20108845

**Authors:** Elisa Benetti, Annarita Giliberti, Arianna Emiliozzi, Floriana Valentino, Laura Bergantini, Chiara Fallerini, Federico Anedda, Sara Amitrano, Edoardo Conticini, Rossella Tita, Miriana d’Alessandro, Francesca Fava, Simona Marcantonio, Margherita Baldassarri, Mirella Bruttini, Maria Antonietta Mazzei, Francesca Montagnani, Marco Mandalà, Elena Bargagli, Simone Furini, GEN-COVID Multicenter Study, Alessandra Renieri, Francesca Mari

## Abstract

Clinical and molecular characterization by Whole Exome Sequencing (WES) is reported in 35 COVID-19 patients attending the University Hospital in Siena, Italy, from April 7 to May 7, 2020. Eighty percent of patients required respiratory assistance, half of them being on mechanical ventilation. Fiftyone percent had hepatic involvement and hyposmia was ascertained in 3 patients. Searching for common genes by collapsing methods against 150 WES of controls of the Italian population failed to give straightforward statistically significant results with the exception of two genes. This result is not unexpected since we are facing the most challenging common disorder triggered by environmental factors with a strong underlying heritability (50%). The lesson learned from Autism-Spectrum-Disorders prompted us to re-analyse the cohort treating each patient as an independent case, following a Mendelian-like model. We identified for each patient an average of 2.5 pathogenic mutations involved in virus infection susceptibility and pinpointing to one or more rare disorder(s). To our knowledge, this is the first report on WES and COVID-19. Our results suggest a combined model for COVID-19 susceptibility with a number of common susceptibility genes which represent the favorite background in which additional host private mutations may determine disease progression.

## INTRODUCTION

Italy has been the first European Country experiencing the epidemic wave of SARS-CoV-2 infection, with an apparently more severe clinical picture, compared to other countries. Indeed, the case fatality rate has peaked to 14% in Italy, while it remains stable around 5% in China. At the time of the study, 12 May 2020, SARS-CoV-2 positive subjects in Italy have reached the threshold of 200.000 cases [1]. Since the beginning of the epidemic wave, one of the first observations has been a highly heterogeneous phenotypic response to SARS-CoV-2 infection among individuals. Indeed, while most affected subjects show mild symptoms, a subset of patients develops severe pneumonia requiring mechanical ventilation with a 20% of cases requiring hospitalization; 5% of cases admitted to the Intensive Care Unit (ICU), and 6,1% requiring intensive support with ventilators or extracorporeal oxygenation (ECMO) machines [2]. Although patients undergoing ventilatory assistance are often older and are affected by other diseases, like diabetes [3], the existing comorbidities alone do not fully explain the differences in clinical severity. As demonstrated for other viral diseases, the basis of these different outcomes there are host predisposing genetic factors leading to different immunogenicity/cytokine responses as well as specific receptor permissiveness to virus and antiviral defence [4–6]. Similarly, during the study of host genetics in influenza disease, a pattern of genetic markers has been identified which underlies increased susceptibility to a more severe clinical outcome (as reviewed in [7]). This hypothesis is also supported by a recent work reporting 50% heritability of COVID-19 symptoms [8].

The identification of host genetic variants associated with disease severity is of utmost importance to develop both effective treatments, based on a personalized approach,and novel diagnostics. Also, it is expected to be of high relevance in providing guidance for the health care systems and societal organizations. However, nowadays, little is known about the impact of host genome variability on COVID-19 susceptibility and severity.

On March 16^th^, 2020 the University Hospital in Siena launched a study named GEN-COVID with the aim to collect the genomic DNA of 2,000 COVID-19 patients for host genetic analysis. More than 30 different hospitals and community centers throughout Italy joined the study and are providing samples and clinical detailed information of COVID-19 patients. This study is aimed to identify common and rare genetic variants of SARS-CoV-2 infected individuals, using a whole exome sequencing (WES) analysis approach, in order to establish an association between host genetic variants and COVID-19 severity and prognosis.

## RESULTS

### Clinical data

The cohort consists of 35 COVID-19 patients (33 unrelated and 2 sisters) of Caucasian ethnicity, except for one North African and one Hispanic. The mean and median age is 64 years (range 31-98): 11 females (median age 66 years) and 24 males (median age 62 years).

The population is clustered into four qualitative severity groups depending on the respiratory impairment and the need for ventilation (groups 1-4 in **Table 1** and different colors in **Fig. 1**) (see Methods section). In the two most severe groups (groups 1 and 2, including 13 patients) there are 11 males and 2 females, while in the two mildest groups (groups 3 and 4 including 22 patients) males are 13 while females are 9.

**Table 1.**
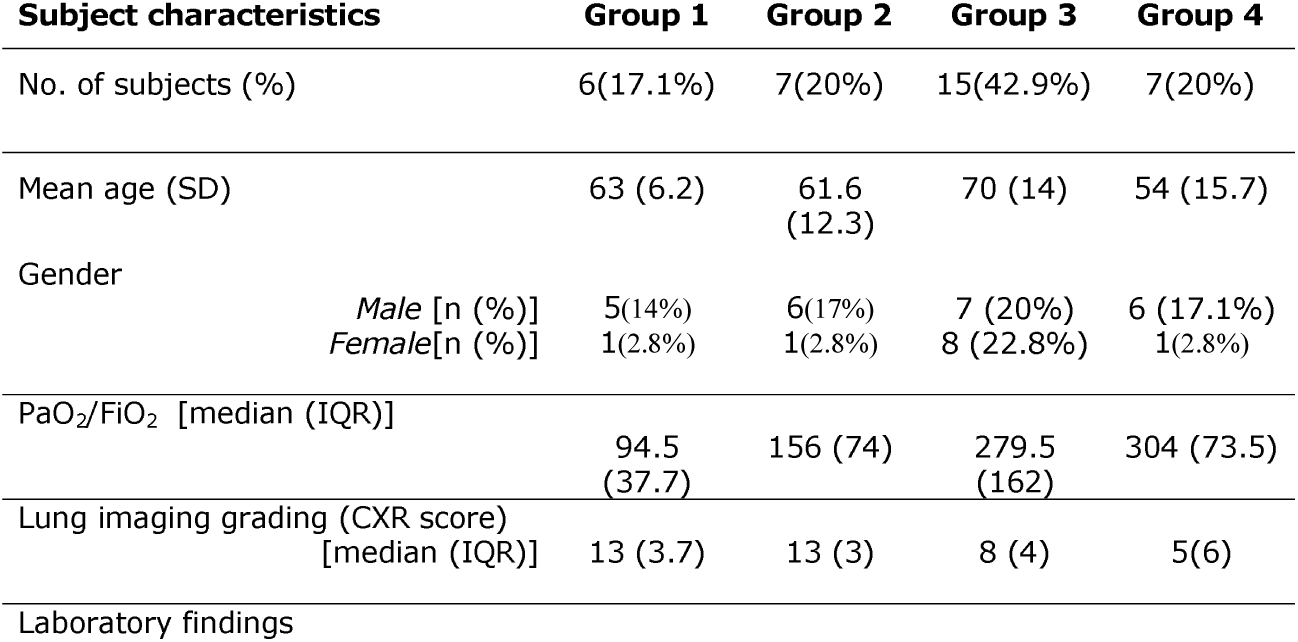
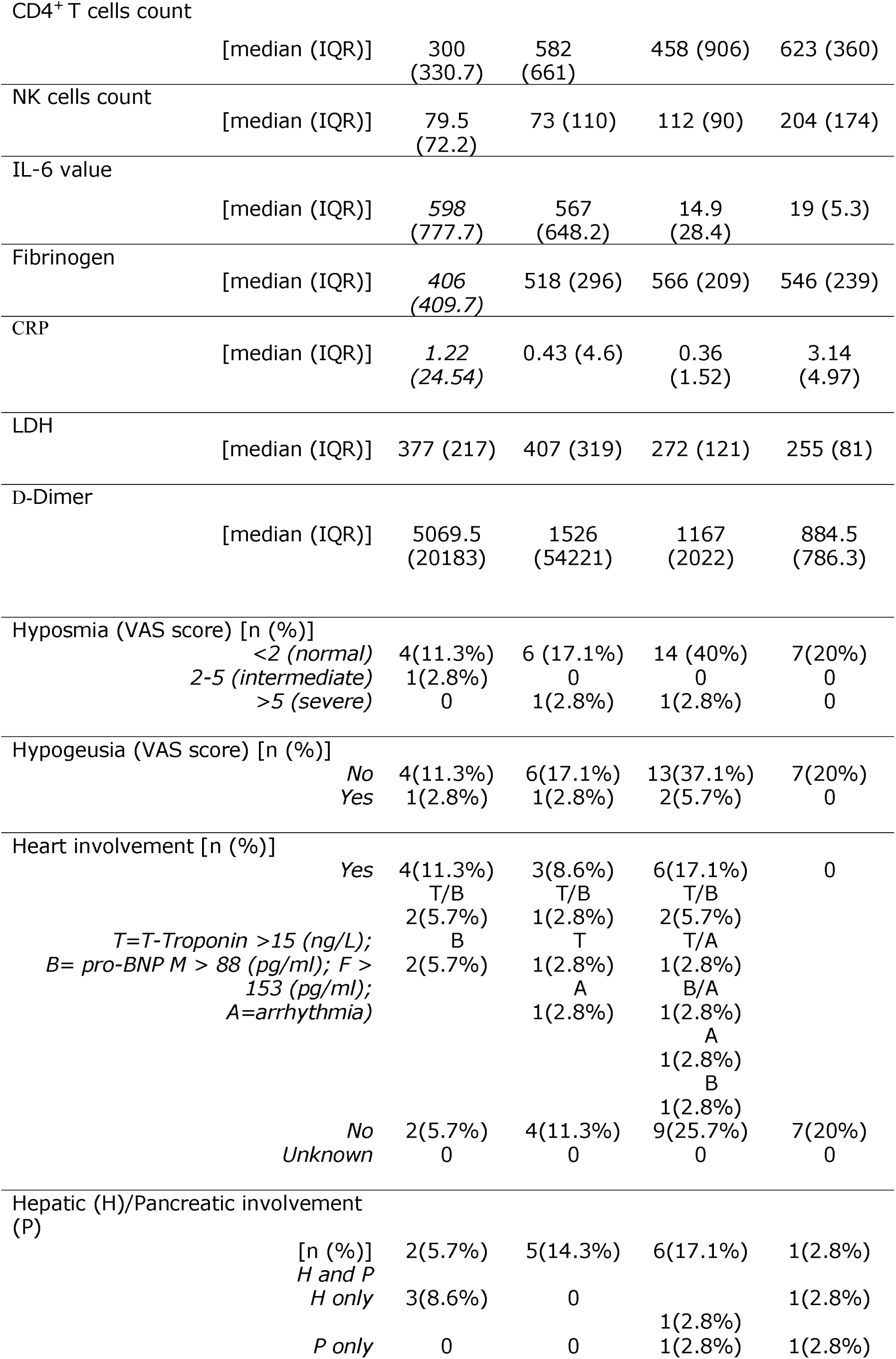
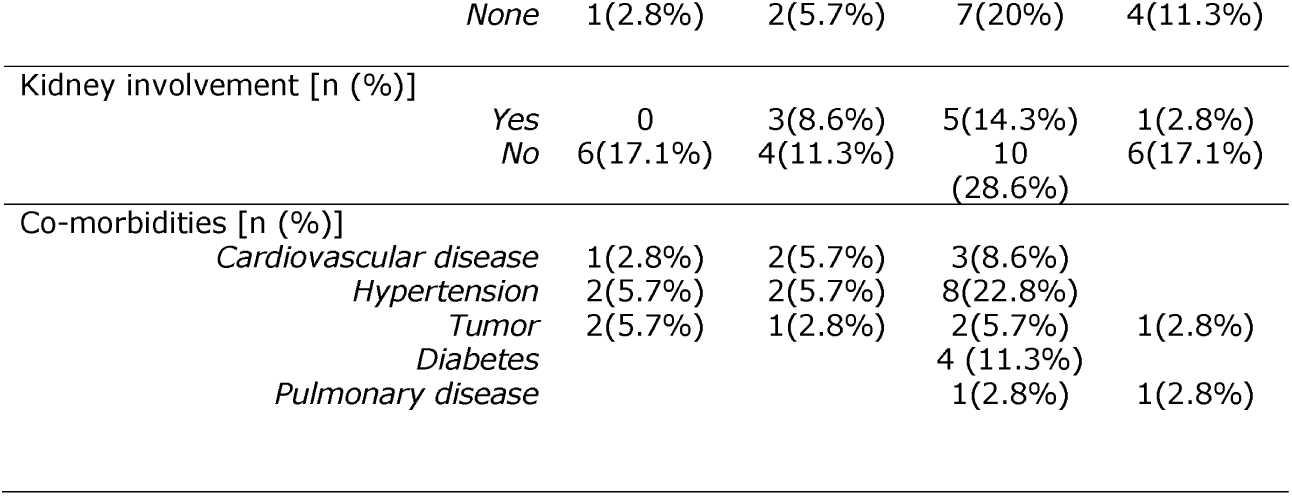
Clinical characteristics COVID19 patients admitted to the University Hospital of Siena (Italy)

**Fig. 1.**
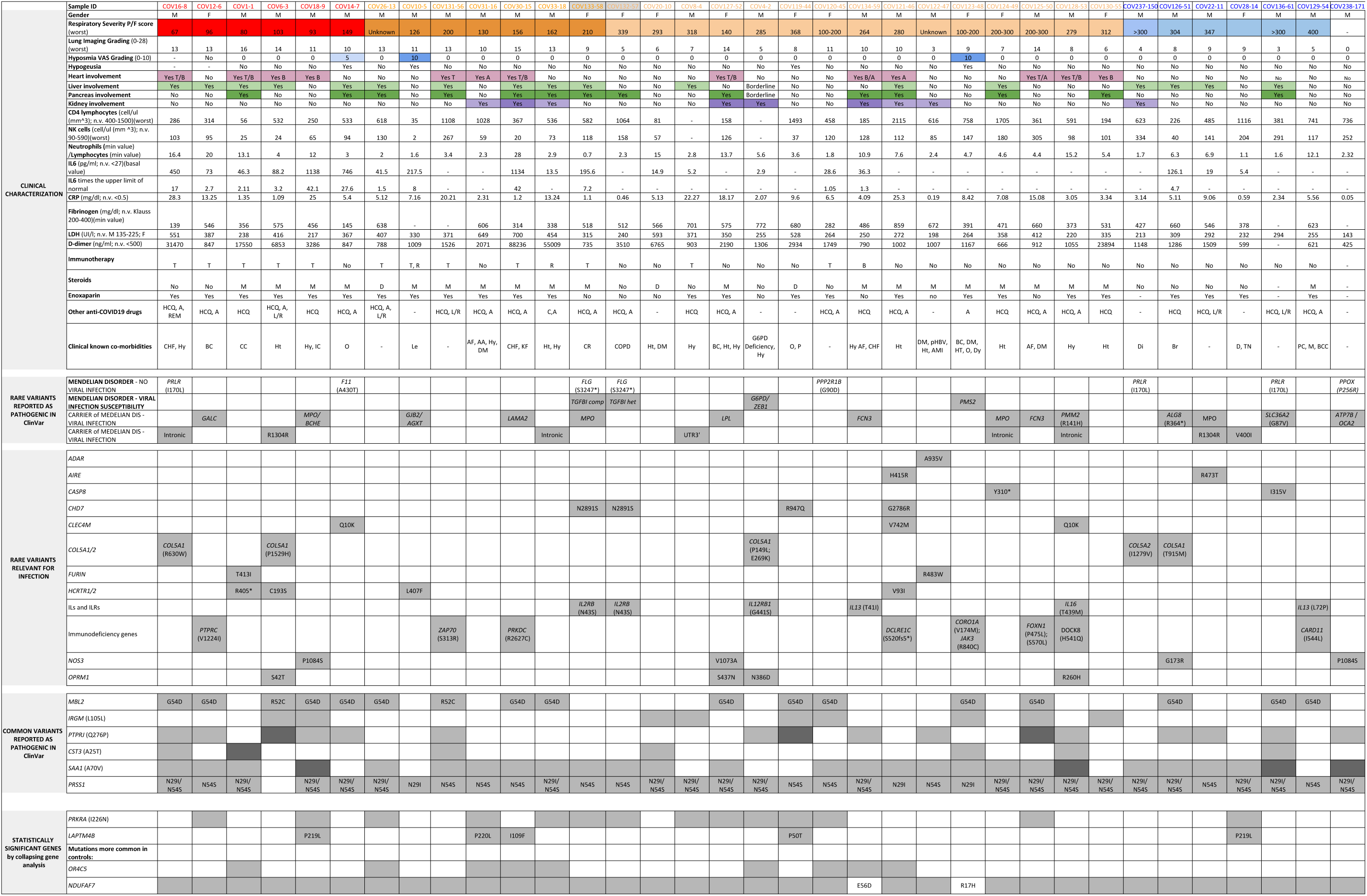
Clinical characteristic and mutated genes. The population is clustered into four qualitative severity groups indicated with different colors depending on the respiratory impairment and the need for ventilation. Red color is used for high care intensity group (those requiring invasive ventilation), orange for intermediate care intensity group (those requiring non invasive ventilation i.e. CPAP and BiPAP, and high-flows oxygen therapy), pink for low care intensity groups (those requiring conventional oxygen therapy) and light blue for very low care intensity groups (those not requiring oxygen therapy). Patients COV132-57 and COV133-58 are reported in grey because they are siblings. A detailed clinical characterization is provided (i.e multiple organs involvement, presence of comorbidity, clinical laboratory parameters, etc.) along with the genetic background for each patient. Liver and pancreas involvement is indicated in green color, heart involvement in magenta and kidney involvement in purple. The presence of hyposmia, classified from 0 to 10 using VAS score, is indicated in light blue. A darker shade of color represents a more severe organ damage. In-silico predicted deleterious variants in genes relevant for infection and pathogenic variants (both common and rare) reported in ClinVar Database are described and a further subdivision between genes involved in a mendelian disorder and/or viral infection susceptibility is provided. For all these gene categories, dark grey is used to identify the homozygous status of the variants while light grey for the heterozygous status. In the end, statistically significant genes obtained after Gene Burden analysis are listed: *PRKRA* and *LAPTM4B* mutational burden resulted to be enriched in the 35 COVID-19 patients compared to the controls, while *OR4C5* and *NDUFAF7* have proven to have an opposite trend, having a mutational burden more enriched in controls. For this reason, for *NDUFAF7, OR4C5*, genes the grey color and the white color are inverted because having less variants and by consequence a more functional gene represents a susceptibility factor. For this category, white color underlies a higher mutational burden while grey color indicates lower mutational burden. Clinical known co-morbidities: CHF (Congestive Heart Failure), Hy (Hypertension), CC (Colon Cancer), Le (Leukemia), DM (Diabetes mellitus), pHBV (Previous HBV infection), AMI (Acute myocardial infarction), BC (Breast Cancer), Ht (Hypothyroidism), IC (Ictus Cerebri), O (Obesity), Dy (Dyslipidemia), KF (Kidney Failure), CR (Congenital Rickets), AF (Atrial Fibrillation), Di (Diverticolosis), Br (Bronchiectasis), COPD (Chronic obstructive pulmonary disease), D (Depression), Tn (Thyroid Nodules), PC (Prostate Cancer), M (Melanoma), BCC (Basal cell carcinoma). Immunotherapy: T (Tocilizumab), R (Ruxolitinib), B (Baricitinib). Steroids: M (Methylprednisolone), D (Dexamethasone). Other anti-COVID19 drugs: HCQ (Hydroxicloroquine), A (Azithromycine), REM (Remdesevir), L/R (Lopinavir/Ritonavir), C (Cloroquine).

Patients were also assigned a lung imaging grading according to X-Rays and CT scans. The mean value is 13 for high care intensity group, 12 for intermediate care intensity group, 8 for low care intensity group and 5 for very low care intensity group.

Regarding immunological findings, a decrease in the total number of peripheral CD4^+^ T cells were identified in 13 subjects, while NK cells’ count was impaired in 10 patients. Six patients showed a reduction of both parameters. IL-6 serum level was elevated in 13 patients.

Hyposmia was present in 3 out of 34 evaluated cases (8.8%), and hypogeusia was present in the same subjects plus another case. These four cases belong to the first three severity groups. Liver involvement was present in 7 cases (20%), while pancreas involvement in 4 cases (11%); 10 patients presented both (29%). Heart involvement was detected in 13 cases (37%). 9 patients (25%) showed kidney involvement. Fibrinogen values below 200 mg/dL were identified in 2 cases (6%), between 200 and 400 mg/dL in 7 cases (20%), and above 400mg/dL in 22 cases (63%). D-dimer value below 500 ng/mL was present in 1 case (3%), between 500 and 5000 ng/mL in 26 cases (74%), and in 7 cases (20%) was 10 times higher than the normal value (>5000 ng/mL) (**Table 1**).

### Unbiased collapsing gene analysis

At first, we tested the hypothesis that susceptibility could be due to one or more common factor(s) in the cohort of patients compared to controls. According to this idea, damaging variants of that/those gene(s) should be either over- or under-represented in patients vs controls. We used, as controls, individuals of the Italian population assuming that the majority of them, if infected, would have shown no severe symptoms. WES data of 35 patients were compared with those of 150 controls (the Siena cohort of the Network of Italian Genomes NIG: http://www.nig.cineca.it) using a gene burden test which compares the rate of disrupting mutations per gene. The variants were collapsed on a gene-by-gene basis, in order to identify genes with mutational burden statistically different between COVID-19 samples and controls. The analysis identified genes harboring deleterious mutations (according to the DANN score) with a statistically significant higher frequency in controls than in COVID-19 patients such as the olfactory receptor gene *OR4C5* (adjusted p-value of 1.5E-10), (**Fig. 2** and **S1 Table**) and *NDUFAF7*, although to a lesser extent (**Fig. 2** and **S1 Table**). For all these genes, the susceptibility factor is represented by the functioning (or more functioning) gene. We also identified two additional genes, *PRKRA* and *LAPTM4B*, for which the probability of observing a deleterious variant was computed higher in the COVID-19 samples compared to controls **(Fig. 2** and **S2 Table)**. In these latter cases, the functioning gene represents indeed a protective factor.

**Fig. 2.**
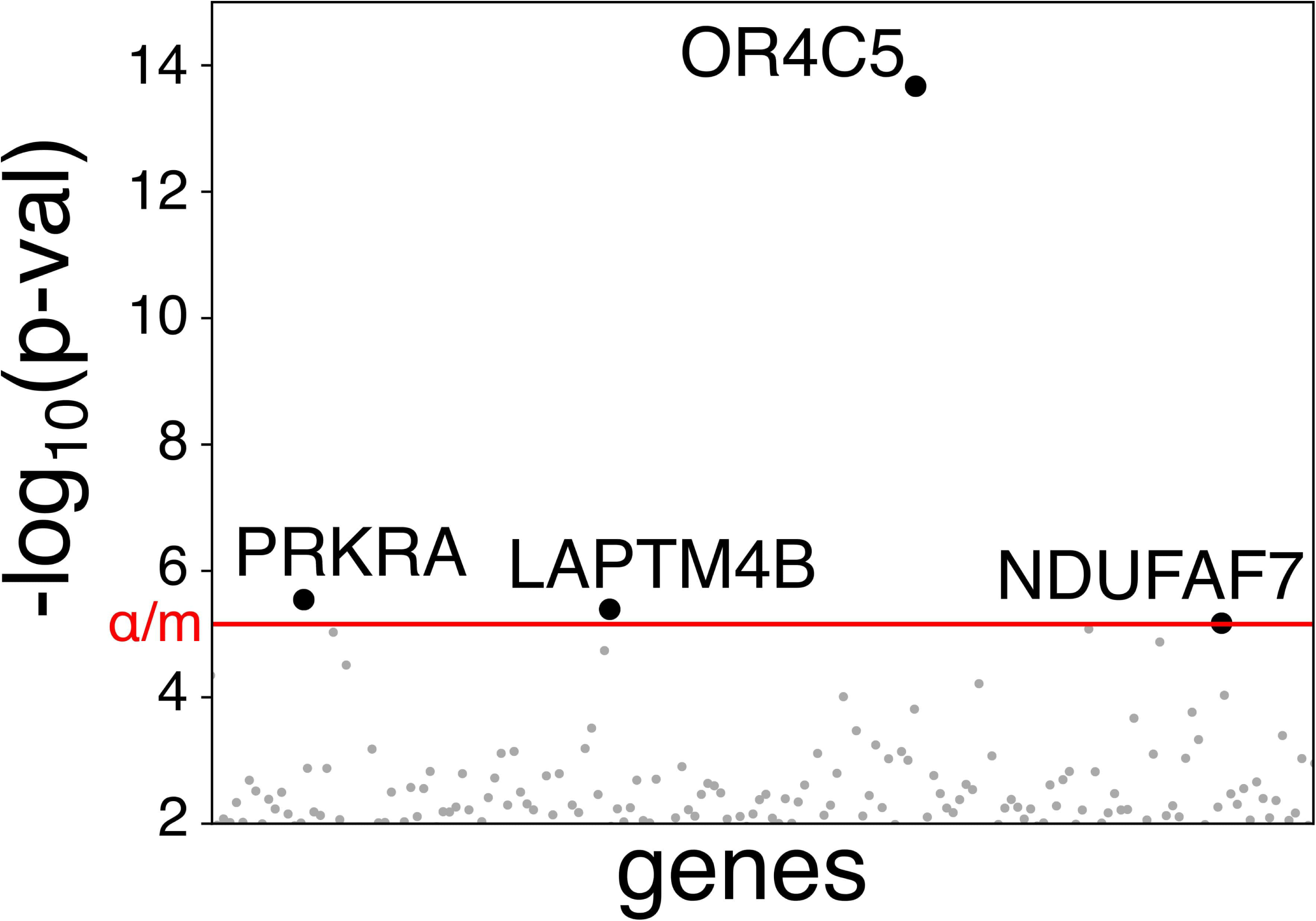
Mutational burden at the gene level. The significance threshold including the Bonferroni correction is shown as a red line (□ = 0.05, number of test, m = 7196). P-values for genes below the significance threshold are shown as grey dots. Black circles are used for the 4 genes that were identified as statistically different between COVID-19 samples and controls.

### Gene analysis using the Mendelian-like model

We then tested the hypothesis that COVID-19 susceptibility is due to different variants in different individuals. A recently acquired knowledge on the genetic bases of Autism Spectrum Disorders suggests that a common disorder could be the sum of many different rare disorders and this genetic landscape can appear indistinguishable at the clinical level [9]. Therefore, we analyzed our cohort treating each patient as an independent case, following a Mendelian-like model. According to the “pathogenic” definition in ClinVar database (https://www.ncbi.nlm.nih.gov/clinvar/), for each patient, we identified an average of 1 mutated gene involved in viral infection susceptibility and pinpointing to one or more rare disorder(s) or a carrier status of rare disorders **(Fig. 1)**. Following the pipeline used in routine clinical practice for WES analysis in rare disorders we then moved forward checking for rare variants “predicted’’ to be relevant for infection by the means of common annotation tools. We thus identified an average of additional 1-5 variants per patient which summed up to the previous identified pathogenic variants **(Fig. 1, S3 Table)**.

### Known common susceptibility/protective variants analysis

We then checked the cohort for known non rare variants classified as either “pathogenic” or “protective” in ClinVar database and related to viral infection. Variants in six different genes matched the term of “viral infection” and “pathogenic” according to ClinVar **(Fig. 1)**. Overall, a mean of 3 genes with “pathogenic” common variants involved in viral infection susceptibility were present **(Fig. 1)**.

Among the common protective variants, we list as example three variants which confer protection to Human Immunodeficiency Virus (HIV), the first two, and leprosy, the third one: a *CCR2* variant (rs1799864) identified in 8 patients, a *CCR5* (rs1800940) in one patient and a *TLR1* variant (rs5743618) in 26 patients (not shown). A *IL4R* variant (rs1805015) associated with HIV slow progression was present in 8 patients (not shown).

### Candidate gene overview

Although not identified by unbiased collapsing gene analysis a number of obvious candidate genes were specifically analyzed. First, we noticed that SARS-CoV-2 receptor, ACE2 protein is preserved in the cohort, only a silent mutation V749V being present in 2 males and 2 heterozygous females. This is in line with our previous suggestion that either rare variants or polymorphisms may impact infectivity [10]. The *IFITM3* polymorphism (rs12252) was found in heterozygosity in 4 patients as expected by frequency. Eight patients had heterozygous missense mutations in *CFTR* gene reported as VUS/mild variants, 7 / 8 being among the more severely affected patients.

## DISCUSSION

In this study, we present a cohort of 35 COVID-19 patients admitted between April and May 2020 to the University Hospital of Siena who were clinically characterized by a team of 29 MDs belonging to 7 different specialties. As expected, the majority of hospitalized patients are males, confirming previously published data reporting a predominance of males among the most severe COVID-19 affected patients [11]. Lung imaging involvement, evaluated through a modified lung imaging grading system, did not completely correlate with respiratory impairment since among the 13 patients who required mechanical ventilation (group 1 and 2), grading was either moderate (10) or mild (3). In line with our previous data, lymphocyte subset immunophenotyping revealed a decrease in the total number of CD4 and NK cells count, especially in the most severe patients [12]. Laboratory tests revealed a multiple-organ involvement, confirming that COVID-19 is a systemic disease rather than just a lung disorder (**Fig. 1**). We thus propose that only a detailed clinical characterization can allow to disentangle the complex relationship between genes and signs/symptoms.

In order to test the hypothesis that the COVID-19 susceptibility is due to one or more genes in common among patients, we used the gene burden test to compare the rate of disrupting mutations per gene. This test has already been successfully applied to discover susceptibility genes for Respiratory Syncytial Virus infection [13]. We identified 2 genes whose damage represents a susceptibility factor. Mutations in *PRKRA* (protein kinase activator A, alias PACT; OMIM# *603424), a protein kinase activated by viral double-stranded RNA may impair the down-stream IFN-mediated immune response [14,15]. Mutations in *LAPTM4B* (Lysosomal Protein Transmembrane 4 Beta) gene, may impair endosomal network, eventually compromising productive viral infection [16,17].

We then identified 2 genes whose damage represents a protective factor: *OR4C5* and *NDUFAF7. OR4C5* is a “resurrected” pseudogene, known to be non functioning in half of the European population, with a frequency of inactive allele of 0.62 in Asians, 0.48 in Europeans and 0.16 in Africans [18,19]. Expression of the “resurrected” pseudogene *OR4C5* may help in triggering the natural immunity leading to virus and cell death [20,21]. It is interesting to note that protein atlas shows *OR4C5* protein expression in the liver without the corresponding mRNA expression (www.proteinatlas.org) suggesting that *OR4C5* reaches the liver through nerve terminals [22]. If this is the case, those individuals expressing the resurrected *OR4C5* gene may have more triggers of innate immunity and subsequently higher liver damage, in agreement with the putative expression of *OR4C5* (white boxes) in patients with liver impairment (**Fig. 1**).

Previous studies reported a prevalence of olfactory disorders in COVID-19 population ranging from 5% to 98%. A recent meta-analysis of 10 studies demonstrated a 52.73% prevalence for smell dysfunction in COVID-19 subjects [23]. In our population, only 3/35 (8.6%) subjects reported olfactory disorders. Both the limited sample size and the characteristic of the population (severely affected hospitalized subjects) could explain this result. However, a report focusing on smell dysfunction in severely affected hospitalized subjects reported a prevalence of 23.7% among 59 patients [24].

We explored the hypothesis that each patient could have one unique combination of rare pathogenic/highly relevant variants related for different reasons to infection susceptibility [9] (**Fig. 1**): G6PD-deficient cells are more susceptible to several viruses including coronavirus and have down-regulated innate immunity (in line with the observed very low levels of IL-6) (**Fig. 1**) [25]; *ZEB1*-linked corneal dystrophy, known to function in immune cells, and playing an important role in establishing both the effector response and future immunity in response to pathogens [26]; *TGFBI* mutations (associated with corneal dystrophy); *ABCC6* gene mutations (associated with pseudoxanthoma elasticum); likely hypomorphic mutations in *CHD7* or *COL5A1/2* variants, playing a role as modulators of immune cells activity and/or response to infections [27–34]; *ADAR*, involved in viral RNA editing; *CLEC4M*, an alternative receptor for SARS-CoV [35] *HCRTR1/2*, receptors of Hypocretin, important in the regulation of fatigue during infections [36]; *FURIN*, a serine protease that cleaves the SARS-Cov-2 minor capsid protein important for ACE2 contact and viral entry into the host cells [37,38].

Finally, interesting rare variants have been identified in NitricOxide synthase *NOS3* and Opioid receptor *OPRM1*. Opioid ligands may regulate the expression of chemokines and chemokine receptors [39]. NitricOxide (NO), mainly produced by epithelial and white blood cells (iNOS) and to a lesser extent by endothelial cells (eNOS), is able to significantly reduce viral infection and replication of SARS-CoV in normal condition through two distinct mechanisms: impairment of the fusion between the spike protein and its receptor ACE2, and reduction of viral RNA production [40]. Mutations in NO synthase may disrupt one or both the above reported functions and clinical trials are ongoing to evaluate the effectiveness of inhaled NO in COVID-19 patients [41,42].

Several rare variants in Interleukins (*ILs*) and Interleukins receptors (*ILRs*) are found. Interleukins are crucial in modulating immune response against all types of infective agents. The variants reported in this study include different interleukins that are not specifically involved in the defense against virus but are critical in balancing both innate and specific adaptive immune response (**Fig. 1**)

Furthemore, we identified common “pathogenic” variants in genes known to be linked to viral infection, such as *MBL2, IRGM* and *SAA1*, and/or specific organ damage as *PRSS1*. Polymorphisms in *PRSS1*, a serine protease secreted from the pancreas, are associated with autosomal dominant hereditary pancreatitis (OMIM#167800) [43]. Polymorphisms in *MBL2*, a mannose-binding lectin secreted by the liver, cause increased susceptibility to infections, possibly due to a negative impact on the ability to mount an immune response [44,45]. Polymorphisms in *IRGM* may lead to impairment of autophagy which in turn controls innate and adaptive immunity [46,47]. *SAA1*, encoding the serum amyloid A (SAA) protein, is an apolipoprotein reactant, mainly produced by hepatocytes and regulated from inflammatory cytokines. In patients with chronic inflammatory diseases, the SAA cleavage product, Amyloid protein A (AA), is deposited systemically in vital organs including liver, spleen and kidneys, causing amyloidosis [48].

For the last above reported genes and pathogenic variants or predicted variants relevant for infection, a statistically significant difference in variant’s frequency was not found between cases and controls looking at either the single variant or the single gene, as a burden effect of variants. However, as depicted in the overall **Fig. 1**, we could hypothesize a combined model in which common susceptibility genes will sum to less common or private susceptibility variants. A specific combination of these 2 categories may determine type (organotropism) and severity of the disease.

Our observations related to the huge amount of data, both on phenome and genome sides, and represented in Figure 1, could also lay the bases for association rule mining approaches. Artificial intelligence techniques based on pattern recognition may discover an intelligible picture which appears blurred at present.

We know that a possible limitation of this study is the heterogeneity of patients and controls, which are not matched for gender, major comorbidities and other clinical characteristics. For this reason, further analyses in a larger cohort of samples are mandatory in order to test this hypothesis of a combined model for COVID-19 susceptibility with a number of common susceptibility genes which represent the fertile background in which additional private, rare or low frequency mutations confer to the host the most favorable environment for virus growth and organ damage.

## METHODS

### Patients clinical data and Samples collection

The GEN-COVID study was approved by the University Hospital of Siena Ethical Review Board (Prot n. 16929, dated March 16, 2020). Thirty-five patients were recruited. WES data of these 35 patients were compared with those of 150 controls (Italian Genomes NIG http://www.nig.cineca.it). Patients have a mean age of 64 years with a Standard Deviation (SD) of 14.3 while the controls have a mean age of 46 years with a SD of 9.5. The percentage of males (M) and females (F) in patients is 68.5% and 31.4% respectively, while in controls is 51% and 49% respectively. The patients are clustered into four qualitative severity groups depending on the respiratory impairment and the need for ventilation: high care intensity group (those requiring invasive ventilation), intermediate care intensity group (those requiring non invasive ventilation i.e. CPAP and BiPAP, and high-flows oxygen therapy), low care intensity group (those requiring conventional oxygen therapy) and very low care intensity group (those not requiring oxygen therapy) (groups 1-4 in **Table 1** and different colors in **Fig. 1**).

Peripheral blood samples in EDTA-containing tubes and detailed clinical data were collected. All these data were inserted in a section dedicated to COVID-19 of the established and certified Biobank and Registry of the Medical Genetics Unit of the Hospital. An example of the Clinical questionnaire is illustrated in **S1 Fig**..

Each patient was assigned a continuous quantitative respiratory score, the PaO2/FiO2 ratio (normal values >300) (P/F), as the worst value during the hospitalization.

Patients were also assigned a lung imaging grading according to X-Rays and CT scans. In particular, lung involvement was scored through imaging at the time of admission and during hospitalization (worst score), annotating the chest X-Ray (CXR) score (in 34 patients) and CT score in 1 patient for whom X-Rays were not available. To obtain the score (from 0 to 28) each CXR was divided in four quadrant (right upper, right lower, left upper and left lower) and for each quadrant the presence of consolidation (0= no consolidation; 1 <50%, 2>50%), ground glass opacities (GGOs: 0= no GGOs, 1<50%, 2 >50%), reticulation (0= no GGOs, 1<50%, 2 >50%) and pleural effusion on left or right side (0= no, 1= minimal; 2= large) were recorded. The same score was applied for CT (1 patient). For each patient, the presence of hyposmia and hypogeusia was also investigated through otolaryngology examination, Burghart sniffin’ sticks [49] and a visual analog scale (VAS). Whenever the sign was present, a score ranging from 0 to 10 was assigned to each patient using VAS where 0 means the best sense of smell and 10 represents the absence of smell sensation [50].

The presence of hepatic involvement was defined on the basis of a clear hepatic enzymes elevation as glutamic pyruvic transaminase (ALT) and glutamic oxaloacetic transaminase (AST) both higher than 40 UI/l. Pancreatic involvement was considered on the basis of an increase of pancreatic enzymes as pancreatic amylase higher than 53 UI/l and lipase higher than 60UI/l. Heart involvement was defined on the basis of one or more of the following abnormal data: Troponin T (>15 ng/L), indicative of ischemic disorder; NT-proBNP (M >88; F >153 pg/ml), indicative of heart failure and arrhythmias (indicative of electric disorder). Kidney involvement was defined in the presence of a creatinine value higher than 1,20 mg/dl in males and higher than 1,10 mg/dl in females (**Fig. 1**).

### Whole Exome Sequencing analysis

Genomic DNA was extracted from peripheral blood using the MagCore®Genomic DNA Whole Blood kit (RBC Biosciences) according to manufacturer’s protocol. Whole exome sequencing analysis was performed on Illumina NovaSeq 6000 system (Illumina, San Diego, CA, USA). DNA fragments were hybridized and captured by Illumina Exome Panel (Illumina) according to manufacturer’s protocol. The libraries were tested for enrichment by qPCR, and the size distribution and concentration were determined using an Agilent Bioanalyzer 2100 (Agilent Technologies, Santa Clara, CA, USA). The Novaseq 6000 platform (Illumina), along with 150 bp paired-end reads, was used for sequencing of DNA.

### Genetic data analysis

Reads were mapped to the hg19 reference genome by the Burrow-Wheeler aligner BWA [51]. Variants calling was performed according to the GATK4 best practice guidelines [52]. Namely, duplicates were first removed by *MarkDuplicates*, and base qualities were recalibrated using *BaseRecalibration* and *ApplyBQSR. HaplotypeCaller* was used to calculate Genomic VCF files for each sample, which were then used for multi-sample calling by *GenomicDBImport* and *GenotypeGVCF*. In order to improve the specificity-sensitivity balance, variants quality scores were calculated by *VariantRecalibrator* and *ApplyVQSR*. Variants were annotated by ANNOVAR [53], and with the number of articles answering the query “gene_name AND viral infection” in Pubmed, where gene_name is the name of the gene affected by the variant.

In order to identify candidate genes according to the Mendelian-like model, rare variants were filtered by a prioritization approach. We used the ExAC database (http://exac.broadinstitute.org/), in particular the ExAC_NFE reported frequency to filter variants according to a minor allele frequency < 0.01. Synonymous, intronic and non-coding variants were excluded from the analysis. Mutation disease database ClinVar (ncbi.nlm.nih.gov/clinvar/) was used to identify previous pathogenicity classifications and variants reported as likely benign/benign were discarded. Filtering and prioritization of variants was completed using the CADD_Phred pathogenicity prediction tool. Finally, we selected genes involved in infection susceptibility using the term “viral infection” as Pubmed database search.

In order to identify genes with a different prevalence of functionally relevant variants between COVID-19 patients and control samples, the following score was calculated:

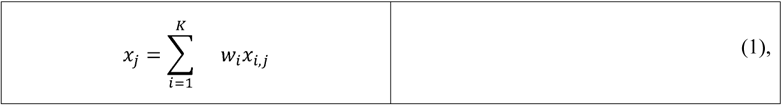

Where *w*_*i*_ is a weight associated with the *i-th* variant; and *x*_*i,j*_, is equal to 0 if the variant is not present in sample *j*, 1 if sample *j* has the variant in heterozygous state, and 2 if sample *j* has the variant is homozygous state. The weight was assumed equal to the DANN score of the variant [54], which provides an estimate of the likelihood that the variant has deleterious functional effects (i.e. variants more likely to have a functional effect contribute more to the score). The sum in equation (1) was performed over all the variants in the gene where the DANN score was available. Genes with less than 5 annotated variants were discarded from the analysis. The scores calculated by equation (1) were ranked for all the samples, and the sum of the ranking for the COVID-19 samples, named *r*_*COVID*_, was calculated. Then, sample labels were permuted 10.000 times, and these permutations were used to estimate the average value and the standard deviation of *r*_*COVID*_ under the null-hypothesis. The p-value was calculated assuming a normal distribution for the sum of the ranking [55]. Moreover, we performed an additional more stringent quality check of genetic variants in the selected genes in order to remove calling artifacts that skipped the previous quality control.

## Data Availability

I declare the availability of all data referred to in the manuscript

## DATA AVAILABILITY

Data about the gene-based analyses and variants are available as Supplementary Material. Genetic data are available in public database of the Italian genetic variability NIG (http://nigdb.cineca.it/index.php#).

## ACKNOWLEDGEMENTS

This study is part of GEN-COVID, https://sites.google.com/dbm.unisi.it/gen-covid the Italian multicenter study aimed to identify the COVID-19 host genetic bases The *Genetic and COVID-19 Biobank of Siena*, member of BBMRI-IT, of Telethon Network of Genetic Biobanks (project no. GTB18001), of EuroBioBank, and of D-Connect, provided us with specimens. We thank the CINECA consortium for providing computational resources and Network for Italian Genomes NIG http://www.nig.cineca.it. We thank private donors’ support to A.R. (Department of Medical Biotechnologies, University of Siena) for the COVID-19 host genetics research project (D.L n.18 of March 17th 2020).

## GEN-COVID Multicenter Study (composition at May 22, 2020, the representative of the GEN-COVID multicenter study is Prof. Francesca Mari)

Gabriella Doddato^1^, Susanna Croci^1^, Laura Di Sarno^1^, Andrea Tommasi^1,2^, Sergio Daga^1^, Maria Palmieri^1^, Massimiliano Fabbiani^5^, Barbara Rossetti^5^, Giacomo Zanelli^3,5^, Paolo Cameli^6^, David Bennett^6^, Simona Marcantonio^7^, Sabino Scolletta^7^, Federico Franchi^7^, Luca Cantarini^9^, Bruno Frediani^9^, Danilo Tacconi^10^, Chiara Spertilli^10^, Marco Feri^11^, Alice Donati^11^, Raffaele Scala^12^, Luca Guidelli^12^, Agostino Ognibene^13^, Genni Spargi^14^, Marta Corridi^14^, Cesira Nencioni^15^, Leonardo Croci^15^, Gian Piero Caldarelli^16^, Maurizio Spagnesi^17^, Paolo Piacentini^17^, Anna Canaccini^18^, Agnese Verzuri^18^, Valentina Anemoli^18^, Massimo Vaghi^21^, Antonella D’Arminio Monforte^22^, Esther Merlini^22^, Mario Umberto Mondelli^23,24^, Stefania Mantovani^23^, Serena Ludovisi^24^, Massimo Girardis^25^, Sophie Venturelli^25^, Andrea Cossarizza^26^, Andrea Antinori^27^, Alessandra Vergori^27^, Stefano Rusconi^28,29^, Matteo Siano^28,29^, Arianna Gabrieli^29^, Daniela Francisci^30,31^, Elisabetta Schiaroli^30^, Pier Giorgio Scotton^32^, Francesca Andretta^32^, Sandro Panese^33^, Renzo Scaggiante^34^, Saverio Giuseppe Parisi^35^, Francesco Castelli^36^, Maria Eugenia Quiros Roldan^36^, Paola Magro^36^, Cristina Minardi^36^, Matteo Della Monica^37^, Carmelo Piscopo^37^, Mario Capasso^38,39,40^, Massimo Carella^41^, Marco Castori^41^, Giuseppe Merla^41^, Filippo Aucella^42^, Pamela Raggi^43^, Matteo Bassetti^44,45^, Antonio Di Biagio^45^, Maurizio Sanguinetti^46,47^, Luca Masucci^46,47^, Chiara Gabbi^19^, Serafina Valente^18^, Susanna Guerrini^8^, Elisa Frullanti^1^, Ilaria Meloni^1^, Maria Antonietta Mencarelli^2^, Caterina Lo Rizzo^2^, Anna Maria Pinto^2^

10) Department of Specialized and Internal Medicine, Infectious Diseases Unit, San Donato Hospital Arezzo, Italy

11) Department of Emergency, Anesthesia Unit, San Donato Hospital, Arezzo, Italy

12) Department of Specialized and Internal Medicine, Pneumology Unit and UTIP, San Donato Hospital, Arezzo, Italy

13) Clinical Chemical Analysis Laboratory, San Donato Hospital, Arezzo, Italy

14) Department of Emergency, Anesthesia Unit, Misericordia Hospital, Grosseto, Italy

15) Department of Specialized and Internal Medicine, Infectious Diseases Unit, Misericordia Hospital, Grosseto, Italy

16) Clinical Chemical Analysis Laboratory, Misericordia Hospital, Grosseto, Italy

17) Department of Prevention, Azienda USL Toscana Sud Est, Italy

18) Territorial Scientific Technician Department, Azienda USL Toscana Sud Est, Italy

19) Independent Scientist, Milan, Italy

20) Department of Cardiovascular Diseases, University of Siena, Italy

21) Chirurgia Vascolare, Ospedale Maggiore di Crema, Italy

22) Department of Health Sciences, Clinic of Infectious Diseases, ASST Santi Paolo e Carlo, University of Milan, Italy

23) Division of Infectious Diseases and Immunology, Department of Medical Sciences and Infectious Diseases, Pavia, Italy.

24) Department of Internal Medicine and Therapeutics, University of Pavia, Italy

25) Department of Anesthesia and Intensive Care, University of Modena and Reggio Emilia, Modena, Italy

26) Department of Medical and Surgical Sciences for Children and Adults, University of Modena and Reggio Emilia, Modena, Italy

27) HIV/AIDS Department, National Institute for Infectious Diseases, IRCCS, Lazzaro Spallanzani, Rome, Italy

28) III Infectious Diseases Unit, ASST-FBF-Sacco, Milan, Italy

29) Department of Biomedical and Clinical Sciences Luigi Sacco, University of Milan, Milan, Italy

30) Infectious Diseases Clinic, Department of Medicine 2, Azienda Ospedaliera di Perugia and University of Perugia, Santa Maria Hospital, Perugia, Italy

31) Infectious Diseases Clinic, “Santa Maria” Hospital, University of Perugia, Perugia, Italy

32) Department of Infectious Diseases, Treviso Hospital, Local Health Unit 2 Marca Trevigiana, Treviso, Italy

33) Infectious Diseases Department, Ospedale Civile “SS. Giovanni e Paolo”, Venice, Italy

34) Infectious Diseases Clinic, ULSS1, Belluno, Italy

35) Department of Molecular Medicine, University of Padova, Italy

36) Department of Infectious and Tropical Diseases, University of Brescia and ASST Spedali Civili Hospital, Brescia, Italy.

37) Medical Genetics and Laboratory of Medical Genetics Unit, A.O.R.N. “Antonio Cardarelli”, Naples, Italy.

38) Department of Molecular Medicine and Medical Biotechnology, University of Naples Federico II, Naples, Italy.

39) CEINGE Biotecnologie Avanzate, Naples, Italy

40) IRCCS SDN, Naples, Italy.

41) Division of Medical Genetics, Fondazione IRCCS Casa Sollievo della Sofferenza Hospital, San Giovanni Rotondo, Italy.

42) Department of Nephrology and Dialysis, Fondazione IRCCS Casa Sollievo della Sofferenza Hospital, San Giovanni Rotondo, Italy.

43) Department of Medical Sciences, Fondazione IRCCS Casa Sollievo della Sofferenza Hospital, San Giovanni Rotondo, Italy.

44) Department of Health Sciences, University of Genova, Genova, Italy.

45) Infectious Diseases Clinic, Policlinico San Martino Hospital, IRCCS for Cancer Research Genova, Italy.

46) Microbiology, Fondazione Policlinico Universitario Agostino Gemelli IRCCS, Catholic University of Medicine, Rome, Italy.

47) Department of Laboratory Sciences and Infectious Diseases, Fondazione Policlinico Universitario A. Gemelli IRCCS, Rome, Italy.

## AUTHOR CONTRIBUTIONS STATEMENT

A.R. and F.M. designed the project and experiments. F.F, M.B, A.E, F.A, E.C., M.dA., S.M, M.A.M., F.M., M.M., E.B., A.R. and F.M. performed clinical evaluations. A.G., F.V., S.A., L.B., and M.B. carried the experiments. E.B. and S.F performed bioinformatic analyses. R.T., C.F., and E.B. carried out statistical analysis and prepared the tables. E.B., A.R. and F.M. wrote the manuscript. E.B. submitted this paper. All authors reviewed the manuscript.

## ADDITIONAL INFORMATION

The authors declare no competing interests.

**Table 1. Clinical characteristics of Italian COVID19 patients**. COVID-19 cohort is grouped in 4 qualitative severity groups depending on the respiratory impairment and the need of ventilation. Group 1 requires invasive ventilation. Group 2 requires CPAP/BiPAP/high-flows oxygen therapy. Group 3 requires conventional oxygen therapy. Group 4 does not require oxygen therapy. Clinical characteristics are listed and the number of patients are indicated for each of them.

**S1 Table. List of genes conferring COVID-19 susceptibility identified with the gene burden test analysis**. Genes harboring deleterious mutations with statistically significant higher frequency in control than in COVID-19 patients are ordered based on p-value deriving from gene burden test analysis. The p-value adjusted is provided after Bonferroni correction.

**S2 Table. List of COVID-19 protective genes identified with the gene burden test analysis**. Genes harboring deleterious mutations with statistically significant higher frequency in COVID-19 patients than in control are ordered based on p-value deriving from gene burden test analysis. The p-value adjusted is provided after Bonferroni correction.

**S3 Table. Rare variants identified in patients cohort**. Rare variants identified in COVID-19 patients according to the Mendelian-like model are reported (see *Methods section*)

**S1 Fig. Clinical Data Questionnaire**. The Questionnaire includes five different categories of data: Patient personal anamnesis and family history, Diagnostic Information, Laboratory Tests, Therapy and Complications. Clinical data were collected in detail for all COVID-19 patients.

## Notes

### Competing Interest Statement

The authors have declared no competing interest.

### Funding Statement

no fundings

### Author Declarations

The GEN-COVID study was approved by the University Hospital of Siena Ethical Review Board (Prot n. 16929, dated March 16, 2020).

