## Supplementary figures and images for "Clinical and molecular characterization of COVID-19 hospitalized patients"

### S1 Fig.

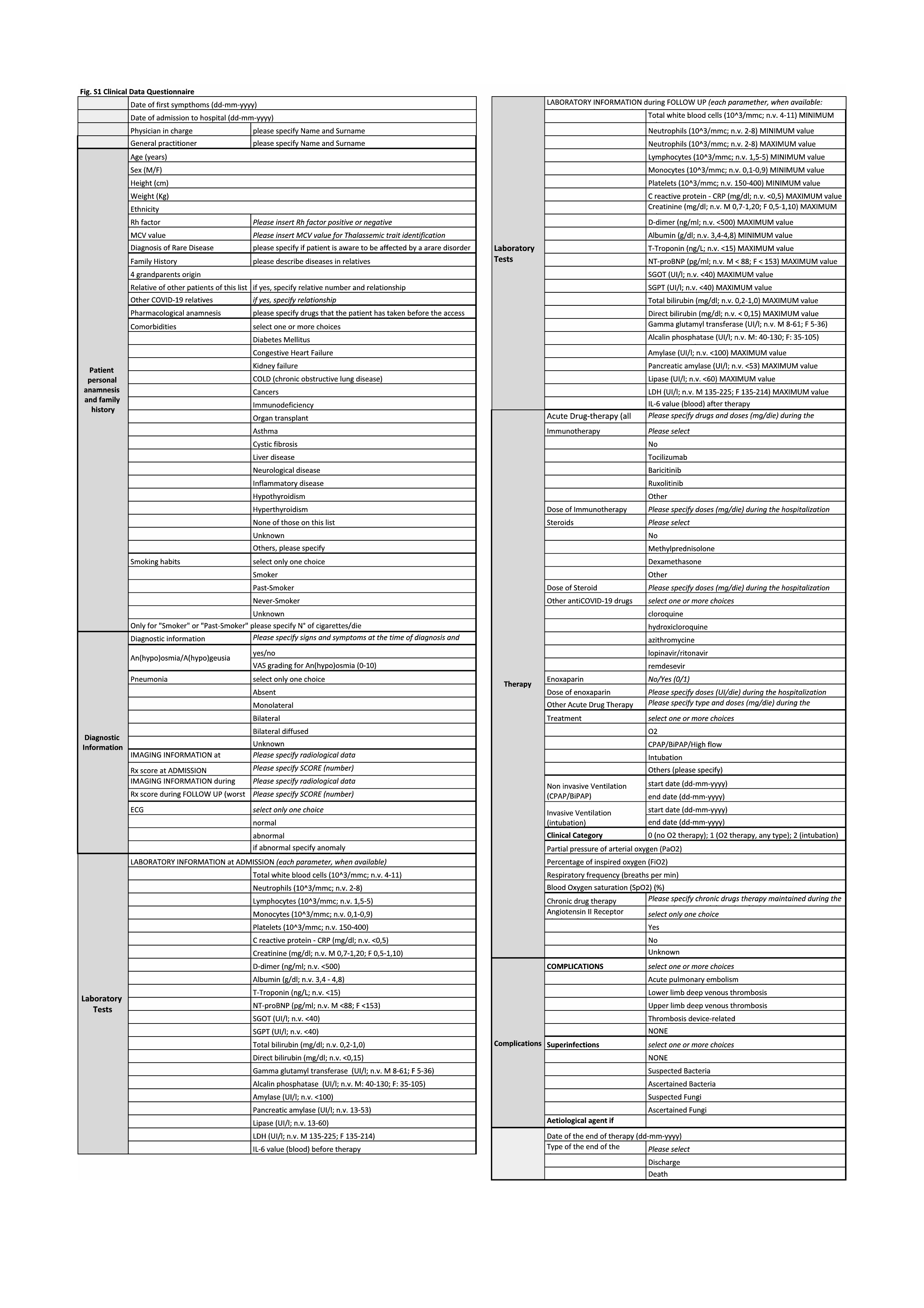
